# Predictors of SARS CoV-2 Infection Among Healthcare Workers: The Impact of Community-Hospital Gradient

**DOI:** 10.1101/2021.05.20.21257518

**Authors:** Fatihan Pınarlık, Zeliha Genç, Süda Tekin, Mahir Kapmaz, Önder Ergönül

**Affiliations:** Koç University School of Medicine, Department of Infectious Diseases and Clinical Microbiology, Istanbul, Turkey; Koç University İşbank Center for Infectious Diseases (KUISCID), Istanbul, Turkey

**Keywords:** Healthcare workers, covid-19, risk factors, seroprevalence, predictor

## Abstract

**Aim:** We aimed to detect the risk factors for SARS-CoV-2 infection among healthcare workers (HCWs) in 2020, before vaccination era.

**Methods:** We surveyed the SARS-CoV-2 infection among the HCWs in a hospital by screening of antibody levels and detection of viral RNA by reverse transcription polymerase chain reaction (RT-PCR) between May 2020 to December 2020. Occupational and non-occupational potential predictors of disease were surveyed for the HCWs included in this study.

**Results:** Among 1925 personnel in the hospital, 1732 were included to the study with the response rate of 90%. Overall seroprevalence was 15% at the end of 2020, before vaccinations started. In multivariate analysis, being janitorial staff (OR:2.24, CI:1.21-4.14, p=0.011), being medical secretary (OR: 4.17, CI: 2.12-8.18, p<0.001), having at least one household member with COVID-19 diagnosis (OR:8.98, CI: 6.64-12.15, p<0.001) and number of household members >3 (OR:1.67, CI:1.26-2.22, p<0.001) were found to be significantly associated with SARS-CoV-2 infection.

**Conclusion:** By the end of 2020, just before the era of vaccination and variants, seroprevalence was 15% among HCWs. Medical secretary and janitorial staff were under increased risk of SARS-CoV-2 infection. Community-hospital gradient can explain the mode of transmission for infection among HCWs. In the setting of this study, community measures were less strict, whereas hospital infection control was adequate and provided necessary personal protective equipment. Increasing risk in larger households and households with diagnosed COVID-19 patient indicates community acquired transmission of the infection.

## Introduction

After its emergence in Wuhan, China in 2019^1^, severe acute respiratory syndrome coronavirus 2 (SARS-CoV-2) spread to world and claimed more than 3 millions of lives up to middle of 2021^2^. Since the beginning of the pandemic, healthcare workers (HCWs) are under increased workload and are facing coronavirus disease 2019 (COVID-19) in the frontlines.

Protection of HCWs from the infection is strategic for the management of the pandemic. Therefore, since the beginning of the pandemic, HCWs were screened for viral RNA and antibody levels to detect the level of infection and also to determine the risk factors among HCWs. In seroprevalence studies, being black^3-9^, being male^8 10 11^, working as frontline worker ^11-15^, working in emergency department^7 9^, intensive care unit^16^, and laboratory^13^ were reported to be associated with higher risk of infection. The availability of PPE is also important because shortage of PPE increases the risk of infection^8^. Some studies reported that non-occupational risk factors such as household contact can increase seropositivity^5 9 17 18^. However, there is no consensus on the risk factors yet.

Detailed and well-designed studies are needed to develop policies for protection of HCWs. By this study, we aimed to determine the level of SARS-CoV-2 infection among HCWs and describe the predictive factors of pre-vaccination era.

## Materials and Methods

### Study Design and Participants

STROBE guideline checklist was used (supplement). All the HCWs in the hospital were aimed to be included in the study. HCWs were reached via e-mails and internal phone calls. Participation was voluntary, and participants were free to leave the study at any time without stating an excuse. All participants were called for antibody level testing at the end of first (May 2020 to end of August 2020) and second wave (September 2020 to end of December 2020) of the pandemic before vaccination and emergence of variants.

Participants with past disease proven by seropositivity or current disease spotted by RT-PCR are put in “infection” group, whereas seronegative and PCR-negative participants are placed in “no infection” group. All participants are surveyed for demographic, occupational and non-occupational risk factors which might be the indicators for SARS-CoV-2 infection.

We screened HCWs for SARS-CoV-2 infection via antibody levels using Elecsys Anti-SARS-CoV (Roche Diagnostics) kits and viral RNA using reverse transcription polymerase chain reaction (RT-PCR) to determine current or past infection. Data collection was terminated by the end of 2020 after the introduction of SARS-CoV-2 vaccines in Turkey.

### Statistical Analysis

All data were collected and stored in a secure database to protect patient confidentiality. Data were analyzed by using Stata 16 computer program. Chi-squared test is used for binary parameters and Mann-Whitney U test is used for continuous variables. Statistically significant risk factors are tested with multivariate analysis and non-significant risk factors are eliminated in stepwise fashion.

### Ethical Approval

The study was approved by Republic of Turkey Ministry of Health (No:2021-04-16T10_28_09) and study was approved by Koç University Institutional Review Board.

## Results

1732 out of 1925 (90%) HCWs responded and volunteered to participate in the study, 67.3% was female and the median age was 28 (min 18 and max 66). By the end of 2020, the rate of seroprevalence was found to be 15% among HCW before the vaccination era.

The comorbidities were reported in 4.4% of the HCWs. Presence of any comorbidity was associated with infection (p = 0.043, Table 1). The professional distribution was dominated by nurses (48.5%) and physicians (11.7%) in the hospital. In univariate analysis, being a medical secretary or janitorial staff are found to be associated with infection (Table 1).

**Table 1.**
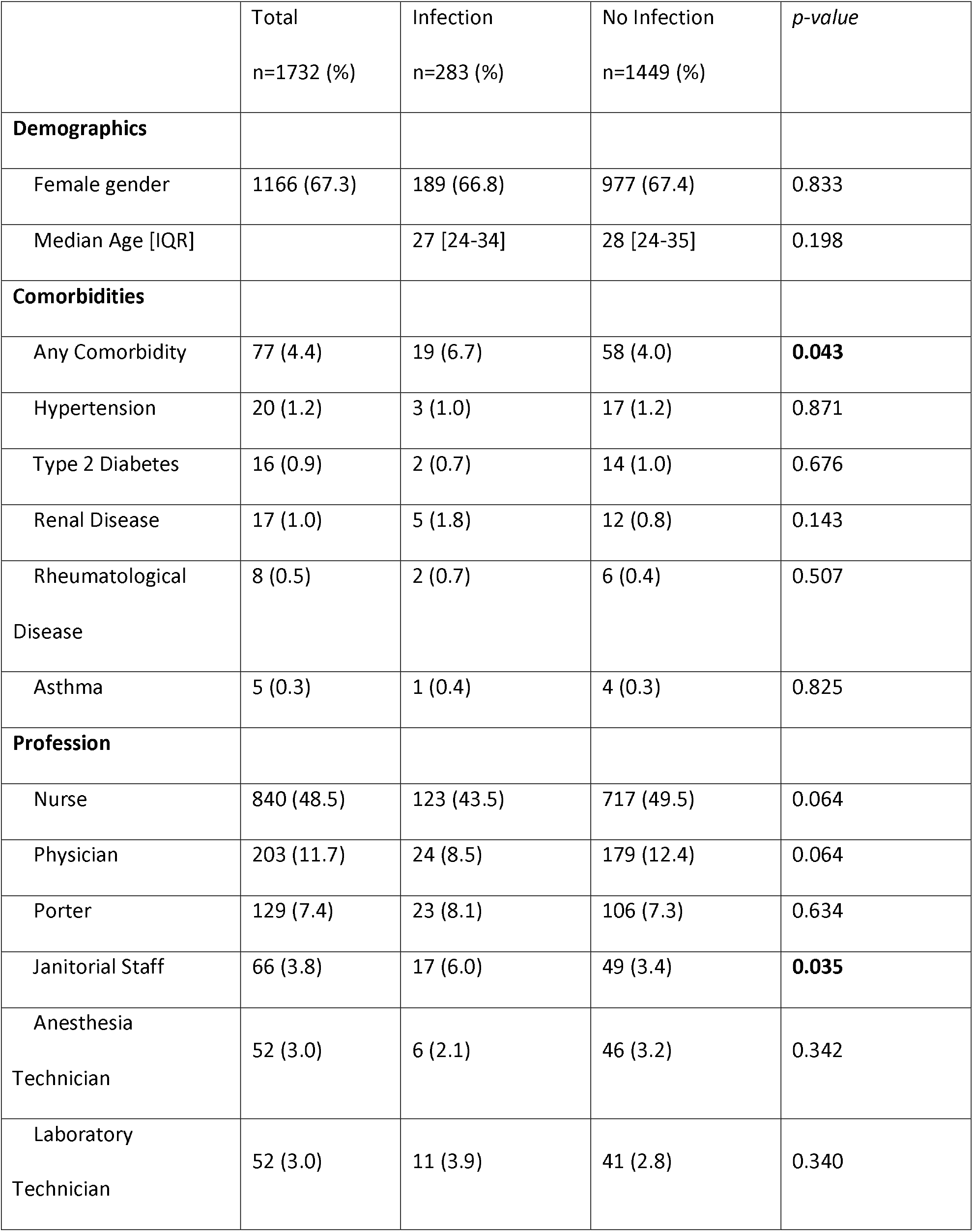
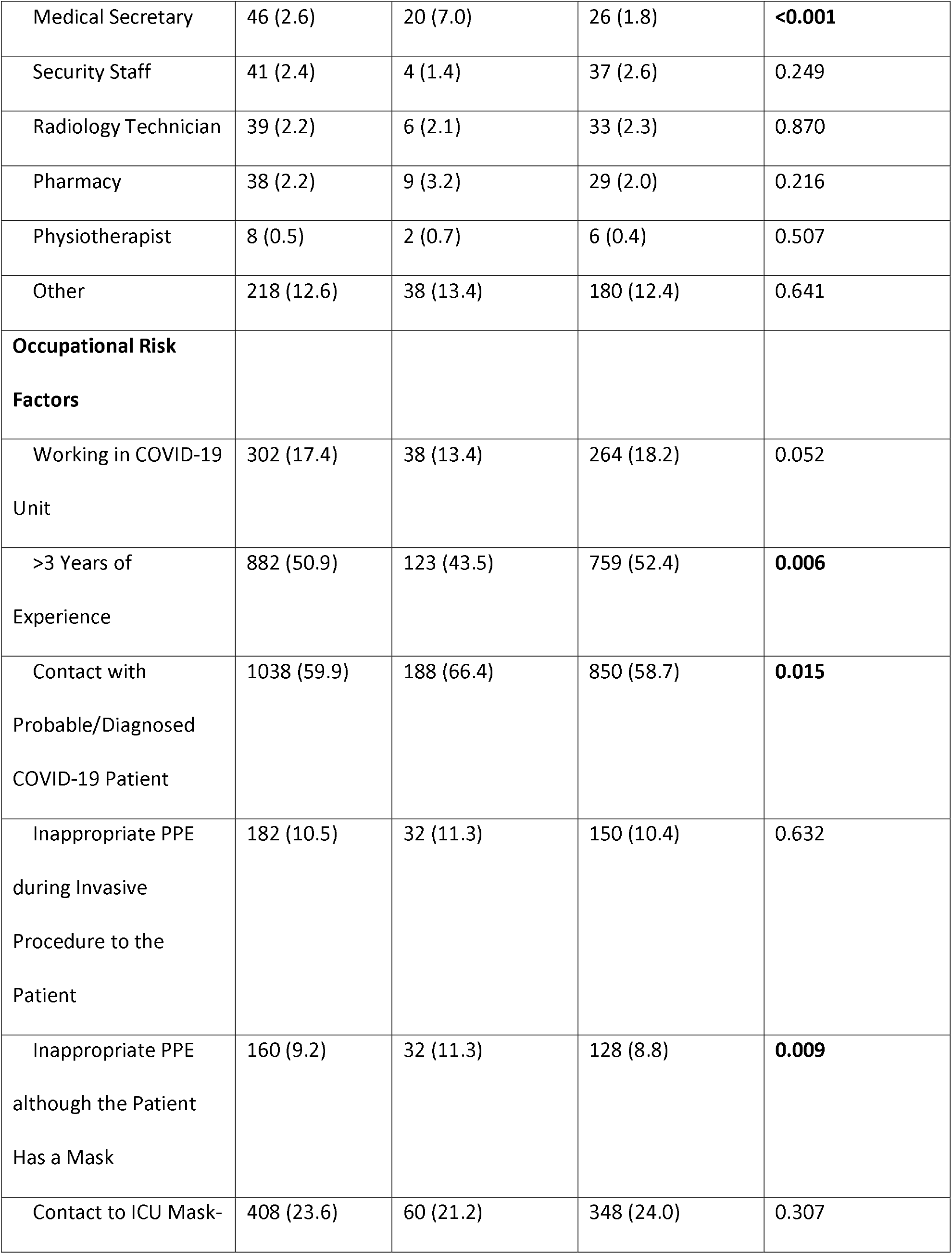
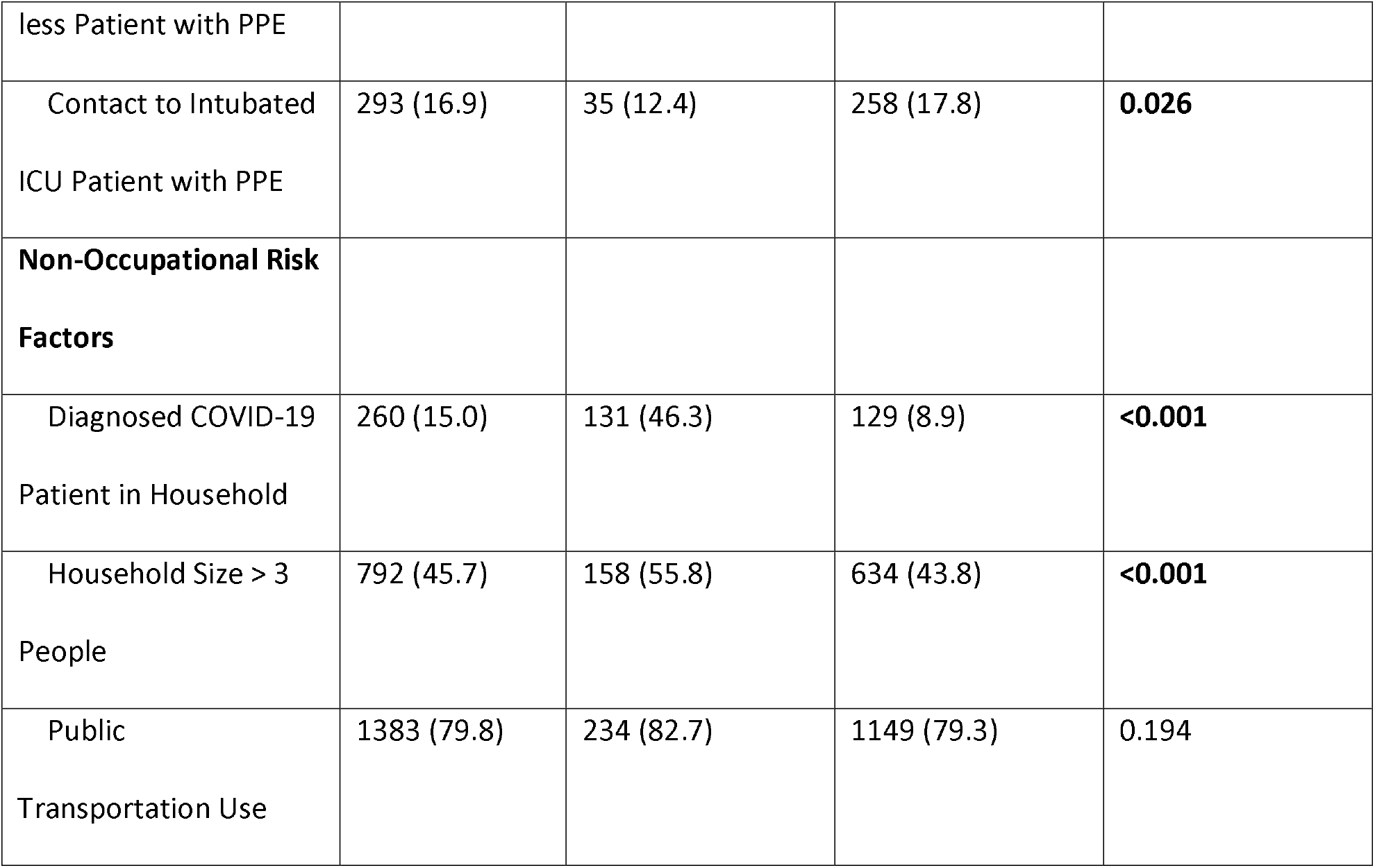
Characteristics and risk factors for SARS-CoV-2 infection

For occupational risk factors, working in the pandemic ward is not associated with infection. However, inappropriate use of PPE by HCWs despite patients wearing masks is associated with increased risk of infection. On the other hand, proper use of PPE in HCW performing intubation is associated with decreased risk of infection. Additionally, having more than three years of experience in the hospital is found to be associated with decreased risk of infection, whereas contact with a COVID-19 patient is associated with increased risk.

There are three non-occupational risk factors investigated in this study. In univariate analysis, using public transportation is not associated with infection (p=0.194). However, presence of diagnosed COVID-19 patient in household and household size are both significantly (p<0.001) associated with infection.

In multivariate analysis, being janitorial staff (OR:2.24, CI:1.21-4.14, p=0.011), being medical secretary (OR: 4.17, CI: 2.12-8.18, p<0.001), having at least one household member with COVID-19 diagnosis (OR:8.98, CI: 6.64-12.15, p<0.001) and number of household members >3 (OR:1.67, CI:1.26-2.22, p<0.001) were found to be significantly associated with SARS-CoV-2 infection (Table 2).

**Table 2:** The predictors of SARS-CoV-2 infection among HCWs.

|  | Univariate analysis |  |  | Multivariate analysis |  |  |
| --- | --- | --- | --- | --- | --- | --- |
| **Risk Factors** | **OR** | **CI** | **p** | **OR** | **CI** | **p** |
| Janitorial Staff | 1.82 | 1.04-3.22 | 0.037 | 2.24 | 1.21-4.14 | 0.011 |
| Medical Secretary | 4.16 | 2.29-7.56 | <0.001 | 4.17 | 2.12-8.18 | <0.001 |
| Diagnosed Patient in Household | 8.82 | 6.56-11.85 | <0.001 | 8.98 | 6.64-12.15 | <0.001 |
| Number of household members > 3 | 1.62 | 1.26-2.10 | <0.001 | 1.67 | 1.26-2.22 | <0.001 |

## Discussion

By this screening study among HCWs that was performed before the era of vaccinations and variants, we investigated the risk factors associated with COVID-19 infection. We detected that being medical secretary (OR: 4.17, CI: 2.12-8.18, p<0.001) or janitorial staff (OR:2.24, CI:1.21-4.14, p=0.011) were associated with increased risk of infection (Table 2). This increase was not caused by the lack of PPE since all HCWs in the hospital were provided with necessary masks, shields, gowns, and other protective equipment where necessary. Informative seminars and posters about SARS-CoV-2 infection were provided to the clinical HCWs, however critical health literacy^19^ might not be achieved among medical secretaries and janitorial staff. In this context, health information clarity should be improved, furthermore, psychological and sociological determinants should be examined for all.

In our study, among the occupational risk factors, contact with COVID-19 patients and inappropriate use of PPE were found to be associated with increased risk of infection. On the other hand, working experience and contact to intubated ICU patient with PPE decreases the risk. These findings suggest that transmission of SARS-CoV-2 can be blocked with appropriate use of PPE. In the settings where PPE are readily available and HCWs are trained against SARS-CoV-2 transmission, occupational transmission could be minimized. We suggest the term “community-hospital gradient” to explain the shift from occupational transmission of SARS-CoV-2 to community acquired form. Community-hospital gradient suggests a more dynamic model and it can be used to express the discrepancy in the literature where some studies^5 9 17 18^ reveal risk factors suggesting transmission from community, whereas others^11-16^ highlights an occupational transmission. Community-hospital gradient favors hospital transmission in the setting of strict community control of SARS-CoV-2 by mask mandates and full lockdowns, along with lack of protective measures in the hospital caused by increased workload and sub-optimal PPE provision. Nevertheless, it can favor community transmission for hospitals where PPE and infection control are provided in the hospital, yet community-level measures are less strict and social setting promotes close contact.

Community-hospital gradient was shifted towards community transmission in the setting of this study and no occupational risk factor was significantly associated with increased infection in multivariate analysis. In contrast, household size and diagnosed patient in the household was associated with increased risk of infection. It should be noted that household size is not studied in the previous literature, although household contact was associated with increased risk by several studies^5 9 17 18^.

Gender and age were studied as demographic risk factors for disease. However, no statistically significant association was found. Some studies in the literature suggest that male gender is a risk factor for SARS-CoV-2 infection^8 10 11^. Although other studies^3-6 9 12-14 17 18 20-25^ have failed to show any significant association which is parallel to our findings. It should be noted that HCWs belong to working population in which the median age and maximum age is lower than the general population. Therefore, the low power of our study, with small sample of older HCWs might not be sufficient to reach the statistical significance.

Similarly, participants with comorbidities constitutes only 4.4% of the studied population showing HCW population studied is healthier than the general population. Although having at least one comorbidity is associated with increased risk of SARS-CoV-2 infection in univariate analysis, low number of participants with chronic disease (77 participants) decreases the capacity to demonstrate any association with each comorbidity, separately. In studies with large sample sizes, some could not show the risk associated with infection^8 11^. On the other hand, Delmas et Al. suggests that diabetes is associated with higher seroprevalence (OR: 1.78, CI: 1.04–3.03)^24^, whereas, Goenka et Al. argues cardiovascular diseases are associated with lower seroprevalence (OR: 0.38, CI: 0.15-0.96)^21^.

Strong part of our study was inclusion of 90% of the HCWs in the hospital to avoid selection bias. On the other hand, there were two main limitations of this study. The first one is the recall bias of participants. We limited recall bias by completing the surveys before serological and PCR testing, and also by using the recording system of occupational health unit. Another limitation is that this study covers only one hospital in Turkey and results may have been affected by hospital and country specific characteristics.

## Conclusion

By the end of 2020, just before the era of vaccination and variants, seroprevalence was 15% among HCWs. Medical secretary and janitorial staff were under increased risk of SARS-CoV-2 infection, because of their exposure in the community or being neglected since they were not in the frontline. Increasing risk in larger households and households with diagnosed COVID-19 patient indicates community acquired transmission of the infection. Community-hospital gradient favors community transmission in case of adequate measures being implemented in the hospital and insufficient community regulations.

## Data Availability

yes

## Acknowledgement

We are thankful to Roche Diagnostics for providing Elecsys Anti-SARS-CoV kits with no charge.

## References

1. Zhu N, Zhang D, Wang W, et al. A Novel Coronavirus from Patients with Pneumonia in China, 2019. The New England journal of medicine 2020;382(8):727–33. doi: 10.1056/NEJMoa2001017

2. Organization WH. WHO Coronavirus (COVID-19) Dashboard: World Health Organization; 2021 [Available from: https://covid19.who.int/ Accessed April 21st 2021.

3. Brant-Zawadzki M, Fridman D, Robinson PA, et al. SARS-CoV-2 antibody prevalence in health care workers: Preliminary report of a single center study. PLoS One 2020;15(11):e0240006. doi: 10.1371/journal.pone.0240006 [published Online First: 2020/11/13]

4. Racine-Brzostek SE, Yang HS, Chadburn A, et al. COVID-19 Viral and Serology Testing in New York City Health Care Workers. American Journal of Clinical Pathology 2020;154(5):592–95. doi: 10.1093/ajcp/aqaa142

5. Martin CA, Patel P, Goss C, et al. Demographic and occupational determinants of anti-SARS-CoV-2 IgG seropositivity in hospital staff. Journal of public health (Oxford, England) 2020 doi: 10.1093/pubmed/fdaa199

6. Jones CR, Hamilton FW, Thompson A, et al. SARS-CoV-2 IgG seroprevalence in healthcare workers and other staff at North Bristol NHS Trust: A sociodemographic analysis. Journal of Infection 2021;82(3):e24–e27. doi: 10.1016/j.jinf.2020.11.036

7. Sydney ER, Kishore P, Laniado I, et al. Antibody evidence of SARS-CoV-2 infection in healthcare workers in the Bronx. Infection control and hospital epidemiology 2020;41(11):1348–49. doi: 10.1017/ice.2020.437

8. Self WH, Tenforde MW, Stubblefield WB, et al. Seroprevalence of SARS-CoV-2 Among Frontline Health Care Personnel in a Multistate Hospital Network - 13 Academic Medical Centers, April-June 2020. MMWR Morbidity and mortality weekly report 2020;69(35):1221–26. doi: 10.15585/mmwr.mm6935e2

9. Eyre DW, Lumley SF, O’Donnell D, et al. Differential occupational risks to healthcare workers from SARS-CoV-2 observed during a prospective observational study. eLife 2020;9 doi: 10.7554/eLife.60675

10. Moscola J, Sembajwe G, Jarrett M, et al. Prevalence of SARS-CoV-2 Antibodies in Health Care Personnel in the New York City Area. JAMA 2020;324(9):893–95. doi: 10.1001/jama.2020.14765

11. Iversen K, Bundgaard H, Hasselbalch RB, et al. Risk of COVID-19 in health-care workers in Denmark: an observational cohort study. The Lancet Infectious Diseases 2020;20(12):1401–08. doi: 10.1016/S1473-3099(20)30589-2

12. Ann-Sofie R, Sebastian H, Anna Mn, et al. SARS-CoV-2 exposure, symptoms and seroprevalence in healthcare workers in Sweden. Nature Communications 2020; 11(1).

13. Andrea C, Valeria G, Teresa E, et al. Risk for SARS-CoV-2 Infection in Healthcare Workers, Turin, Italy. Emerging Infectious Diseases 2021; 27(1).

14. Lidström AK, Sund F, Albinsson B, et al. Work at inpatient care units is associated with an increased risk of SARS-CoV-2 infection; a cross-sectional study of 8679 healthcare workers in Sweden. Upsala journal of medical sciences 2020;125(4):305–10. doi: 10.1080/03009734.2020.1793039

15. Grant JJ, Wilmore SMS, McCann NS, et al. Seroprevalence of SARS-CoV-2 antibodies in healthcare workers at a London NHS Trust. Infection control and hospital epidemiology 2021;42(2):212–14. doi: 10.1017/ice.2020.402

16. Blairon L, Mokrane S, Wilmet A, et al. Large-scale, molecular and serological SARS-CoV-2 screening of healthcare workers in a 4-site public hospital in Belgium after COVID-19 outbreak. Journal of Infection 2021;82(1):159–98. doi: 10.1016/j.jinf.2020.07.033

17. Steensels D, Oris E, Coninx L, et al. Hospital-Wide SARS-CoV-2 Antibody Screening in 3056 Staff in a Tertiary Center in Belgium. JAMA 2020;324(2):195–97. doi: 10.1001/jama.2020.11160

18. Dimcheff DE, Schildhouse RJ, Hausman MS, et al. Seroprevalence of severe acute respiratory syndrome coronavirus-2 (SARS-CoV-2) infection among Veterans Affairs healthcare system employees suggests higher risk of infection when exposed to SARS-CoV-2 outside the work environment. Infection control and hospital epidemiology 2021;42(4):392–98. doi: 10.1017/ice.2020.1220

19. Chinn D. Critical health literacy: A review and critical analysis. Social Science & Medicine 2011;73(1):60–67. doi: 10.1016/j.socscimed.2011.04.004

20. Mina P, Andreas K, Ioanna DP, et al. Antibodies against SARS-CoV-2 among health care workers in a country with low burden of COVID-19. PLoS ONE 2020; 15(12).

21. Goenka M, Afzalpurkar S, Goenka U, et al. Seroprevalence of COVID-19 Amongst Health Care Workers in a Tertiary Care Hospital of a Metropolitan City from India. The Journal of the Association of Physicians of India 2020;68(11):14–19.

22. Jeremias A, Nguyen J, Levine J, et al. Prevalence of SARS-CoV-2 Infection Among Health Care Workers in a Tertiary Community Hospital. JAMA internal medicine 2020 doi: 10.1001/jamainternmed.2020.4214

23. Plebani M, Padoan A, Fedeli U, et al. SARS-CoV-2 serosurvey in health care workers of the Veneto Region. Clinical chemistry and laboratory medicine 2020;58(12):2107–11. doi: 10.1515/cclm-2020-1236

24. Delmas C, Plu-Bureau G, Canoui E, et al. Clinical characteristics and persistence of SARS-CoV-2 IgG antibodies in 4607 French healthcare workers: Comparison with European countries. Infection Control and Hospital Epidemiology 2020 doi: 10.1017/ice.2020.1309

25. Xu X, Sun J, Nie S, et al. Seroprevalence of immunoglobulin M and G antibodies against SARS-CoV-2 in China. Nature medicine 2020;26(8):1193–95. doi: 10.1038/s41591-020-0949-6

